# Cellular, Connectomic, and Cognitive Impact of Glioma and its Surgical Resection

**DOI:** 10.1101/2025.01.27.25320920

**Authors:** A Poologaindran, AI Luppi, MG Hart, T Santarius, S Price, ME Sughrue, J Seidlitz, RAI Bethlehem, M Assem, Y Erez, J Duncan, ET Bullmore, J Suckling, R Romero-Garcia

## Abstract

Awake surgery with intraoperative direct electrical stimulation (DES) is the gold-standard to maximize the extent of resection in diffuse cerebral gliomas (Duffau et al. 2023). While this approach is effective in testing for simple motor and language functions, it is inadequate for mapping higher-order cognitive functions such as attention, working memory, and cognitive control. Given that systems neuroscience is moving away from a localizationist to a connectomic perspective of human brain function, ideally, we could better understand how gliomas integrate within the connectome and how performing surgery on the brain’s mesoscale hub architecture affects long-term cognitive outcomes. To address problem, we combined cellular, connectomic, and cognitive data from healthy individuals (n=629) across the lifespan, cross-sectional glioma imaging (n=98), the Allan Human Brain Atlas (n=6), and a rare cohort of diffuse glioma patients (n=17) followed longitudinally as they underwent neurosurgery. First, we validate that meta-analytic cognitive activation maps co-localize with the Multiple Demand (MD) system and show that diffuse gliomas preferentially localize to the ‘core’ of this brain network. Second, cellular decoding of the MD core network reveals that it is uniquely enriched with oligodendrocyte precursor cells, glioma proto-oncogenes, and 5HT2-serotonergic neurotransmission. Third, the MD system is preferentially enriched for connector hubs to scaffolding the brain’s mesoscale hub architecture and that diffuse gliomas induce reorganization in this architecture thereby minimizing cognitive deficits. Lastly, surgical resection of connector, rather than provincial, hubs leads to long-term cognitive deficits while maintenance or dissolution of interhemispheric modularity predicted long-term cognitive outcomes. With the recent demonstration of the high concordance between DES and functional brain mapping (Saurrubo et al. 2024), this study provides new insight into how gliomas integrate within the connectome and that mapping the mesoscale hub architecture in each patient may improve presurgical mapping and postsurgical rehabilitation. Given the small but deeply sampled neurosurgical cohort, additional studies are now warranted to assess the value of mapping mesoscale connectivity for presurgical mapping and ‘interventional neurorehabilitation’ (Poologaindran et al. 2022).

## Cellular, Connectomic, and Cognitive Impact of Glioma and its Surgical Resection

Diffuse gliomas, which account for approximately 15% of all cerebral gliomas, arise from dysregulated glial precursor cells^1^. These tumours intricately integrate within the brain’s parenchyma thereby minimizing disruption to higher-order cognitive functions such as working memory, attention, and inhibitory control^2,3^. Fortunately, supramaximal surgery – removing tissue beyond the radiologically defined glioma margins – has been demonstrated to increase progression-free survival, reduce the likelihood of malignant transformation, and increase overall survival, irrespective of the tumour’s molecular subtype^5,6^. To achieve maximal extent of resection (EOR), awake surgery with intraoperative direct electrical stimulation (DES) is the gold-standard approach ^7,9,10^. DES enables a functional approach to surgery that enables optimizing “onco-functional balance” –maximal tumour resection while minimizing neurological risk^10^. Awake DES operates on the principle that it is highly sensitive for detecting local cortical areas and intraoperatively testing brain functions. However, while DES is effective in mapping basic motor and language functions, it is limited in its ability to assess higher-order cognitive functions due to the spatially distributed nature of these functions and intraoperative time constraints^11^. A recent meta-analysis of 232 awake intraoperative mapping revealed that while motor and language cortical areas were commonly tested, assessment of higher-order cognitive functions was exceedingly rare^12^. Despite surgery significantly prolonging life, cognitive deficits post-surgery is a significant concern which can severely impact the quality of life in glioma patients and limit their return to employment, fulfill familial duties, and maintain social cognition^7,8^. Ideally, non-invasive functional mapping approaches could be deployed to map brain circuits subserving higher-order cognition; this would complement DES and guide surgical resection approaches to preserve ’onco-functional balance at the individual level^10,14^.

Systems neuroscience has recently embraced a shift away from the ‘localizationist’ to a ‘connectome’ perspective on human brain function^15,17^. It is now widely accepted that higher-order cognition is scaffolded by spatially distributed networks that provide flexible, top-down control of behaviour that is tuned based on a moment-to-moment fluctuations in internal and external demands^15–18^. Specifically, the Multiple Demand (MD) system is a well-characterized frontoparietal brain network causally active during diverse cognitive tasks, including selective attention, working memory, task-switching, novel-problem-solving, among many more^19–22^. Typically, the MD system is mapped using task-based functional mapping paradigms and in non-clinical research settings. Recently, Assem and colleagues presented a highly granular model of the MD system partitioning it into a ‘core’ critical and ‘penumbra’ of supportive networks^23^ (Figure 1). These frontal and parietal MD brain regions have traditionally been considered as redundant in neurosurgical planning not eloquent in the classical Spetzler-Martin grading for arteriovenous malformations which assumes homogeneity among individuals^68,69^. Nevertheless, in neurosurgical settings, patients would encounter challenges completing the extensive battery of tasks required to map the MD system using traditional methods. Additionally, clinical scanning time is more limited and stricter compared to compared to research environments. Thus, it is essential to understand how brain areas that co-activate during cognitive tasks are functionally connected at The relationship between task and task-free functional architecture of the brain has been the subject of intense and rigorous investigation. Cole’s and colleagues seminal work indicates that the brain’s functional network architecture during diverse cognitive tasks (i.e, working memory, selective attention) is primarily shaped by an intrinsic network that is also present at rest (r=0.9 between resting vs. task states)^24,25^. Gratton and colleagues showed that while statistically changes in connectivity occurs in functional connectivity occurs across tasks, likely reflecting task-compliance and strategy, these changes are relatively small compared to the intrinsic architecture present at rest^26^. Moreover, empirical work suggests that task-induced network reconfigurations are modest compared to the intrinsic functional architecture present at rest^24,27^ and that task-induced activity can be well-predicted from resting-state data alone^25,28^. Thus, these results suggest that task-induced reconfigurations occur against a backdrop of stable intrinsic functional networks that support diverse cognitive functions and that mapping the MD system for cognition through a task-free approach may be most practical and suitable for clinical neurosurgery.

**Figure 1.**
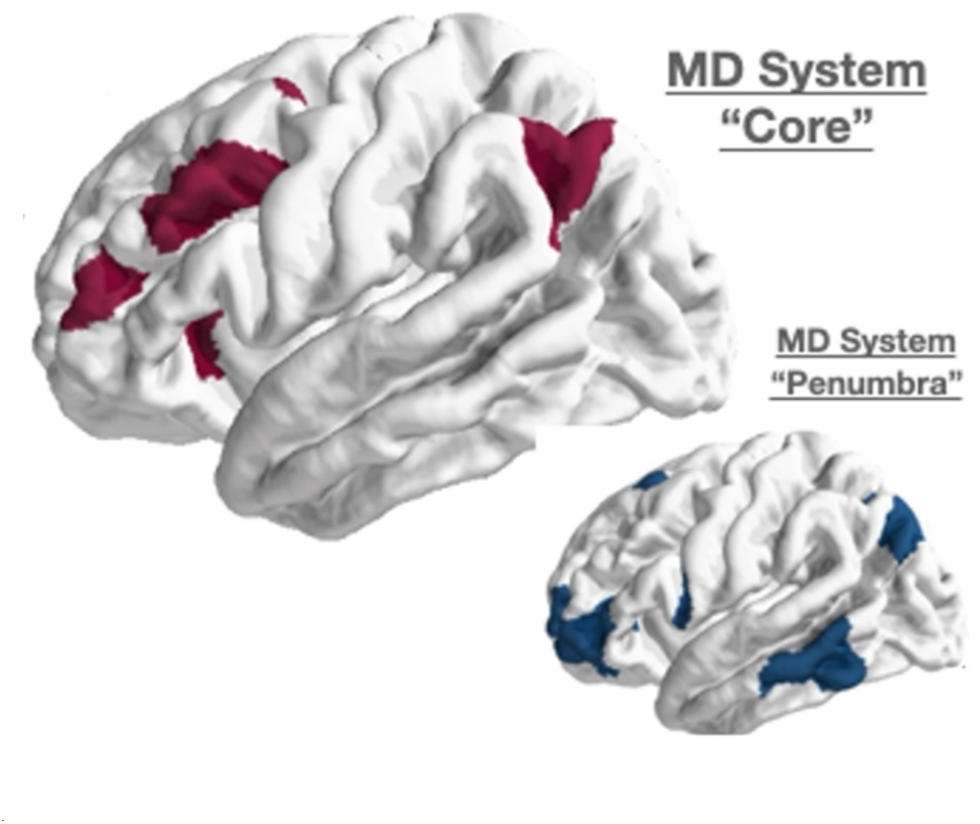
**Top** - Lateral surface of the Multiple Demand (MD) ‘core’ system for domain-general cognition as mapped by Assem and colleagues^23^. Medial surface (not plotted) includes area 8BM (Supplementary and Cingulate Eye Field) **Bottom-** Lateral surface of the MD ‘penumbra’ system rest, as this knowledge would be key to integrating functional mapping into the clinical neurosurgery workflow.

**Figure 1.**
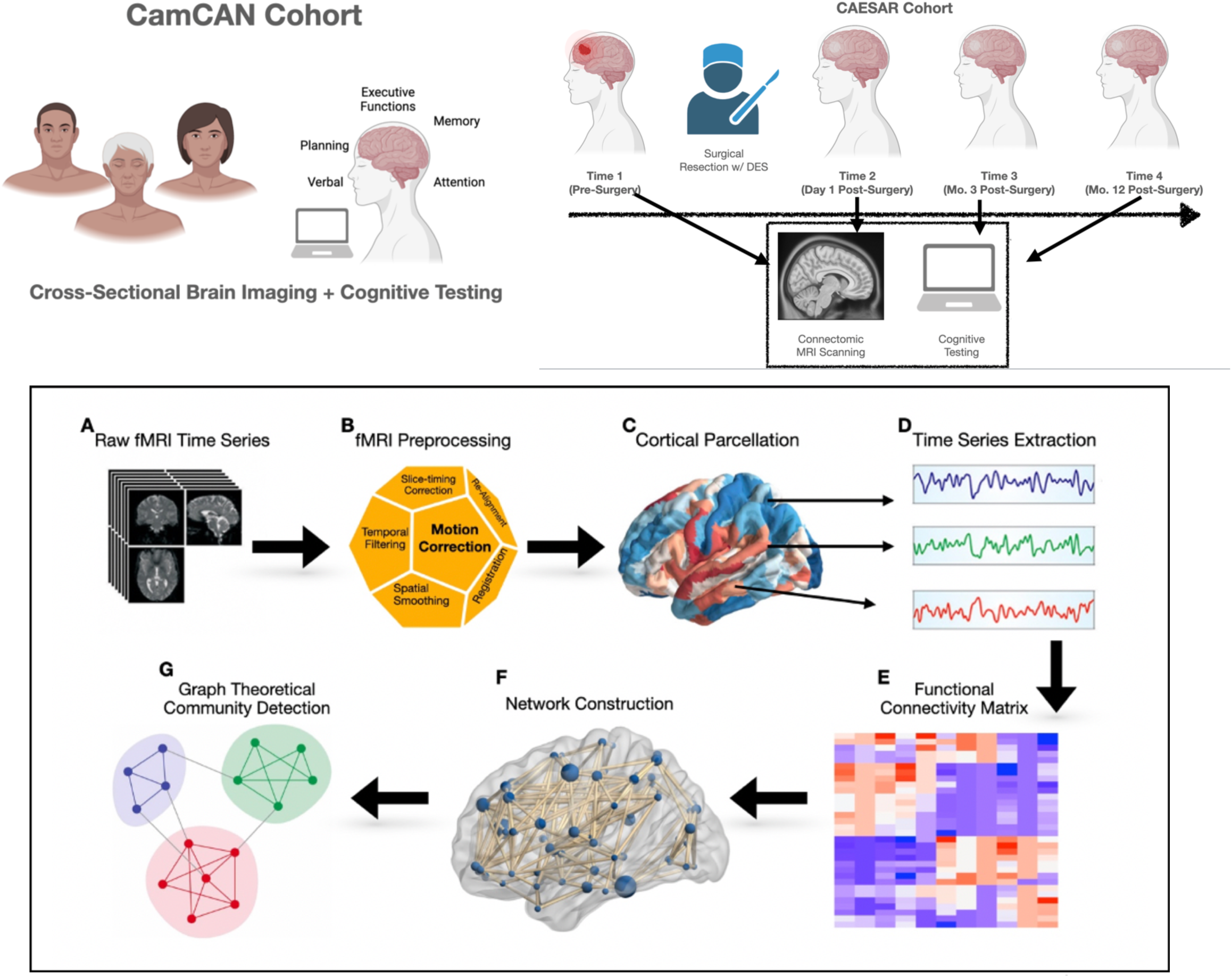
Neurosurgical Study Design and Mapping Mesoscale Cortical Dynamics. Top Panel (used with permission^59^) Participants were recruited by a multidisciplinary neuro-oncology team based on their eligibility for surgical resection. Data collection involved resting-state functional MRI (rs-fMRI) and cognitive assessments conducted at several time points: pre-surgery, immediately post-surgery (<24 hours), and at 3 and 12 months post-surgery. **Bottom Panel:** A: rs-fMRI was obtained from all participants at each time point. B: Data underwent several pre-processing steps described in text. C: After registration to to anatomical space, each patient’s cortex underwent Glasser 360 regional parcellation. D. The BOLD time series was extracted for each parcel. E. Functional connectivity (FC) was estimated by correlating the time series data between all parcels. F. A brain network was constructed using the FC matrix and all 360 regions. G. The Louvain community detection algorithm was applied to the FC matrix to evaluate modularity and the community structure of the network..

With the advent of connectomics^17^, the brain can be modelled as a graph to create a macroscale ‘wiring’ diagram. Like social networks, the brain’s architecture is driven by modularity which quantifies the extent of network integration and segregation; networks with high modularity have dense within module connections and sparser connections between modules^29–32^. The brain regions that link two spatially distributed networks are called “connector hubs” and serve to integrate information between different networks as they integrate control functions and enable higher-order cognition^29^. Moreover, a modular brain architecture enables anatomically segregated and functional specialized communities to reduces the network’s overall wiring and metabolic costs^30^. Crucially, higher levels of brain modularity are predictive of intervention-related cognitive gains across several interventions (i.e., cognitive training, exercise) and populations (i.e., healthy ageing adults, stroke, and traumatic brain injury)^33,34^. Collectively, mapping the modular or mesoscale architecture of the brain provides a representation of global and local scale neural dynamics, and identifying neurosurgical patients susceptible of modular breakdown during surgical planning would be of clinical importance to avoid long-term post-surgical deficits. Recently, Moretto and colleagues demonstrated that functional brain networks mapped via task-free functional MRI has a remarkable consistency (∼5 mm) with the ’ground-truth’ DES approach ^35^. Thus, to complement DES, functional mapping of the brain’s modular (mesoscale) architecture and connector hubs represents a promising approach to map higher-order cognition in neurosurgical patients.

In this study, we performed a series of inter-related investigations to explore the interplay between the MD system, cognition, and glioma resection. We first we sought to elucidate a neurobiological basis to how the brain can paradoxically support both cognition and gliomagenesis. We then further examined the relationship between the MD system and mesoscale hub architecture in healthy individuals across the lifespan and investigated how glioma infiltration of the MD system impacts cognitive outcomes and hub plasticity. Finally, we conducted a prospective longitudinal study in a rare neurosurgical cohort to determine how diffuse gliomas integrate within the brain’s hub architecture and how neurosurgical resection affects hub topology and long-term cognitive outcomes. Given that the brain’s mesoscale architecture has been extensively studied in various neurological, psychiatric, and developmental contexts, for the first time, we sought to investigate these concepts in neurosurgical patients by investigating how glioma infiltration and surgical resection of critical brain hubs affect the brain’s modular organization and long-term cognitive outcomes.

## Methods

### Section A Linking the Multiple Demand (MD) System to Cognitive Activation and Glioma Vulnerability Maps

In the first stage of our endeavour, we sought to better understand how the MD system spatially relates to cognitive activation maps and glioma vulnerable brain regions. We then aimed to interrogate the underlying neurobiology of the MD system to explain how it may paradoxically harbour to both cognition and glioma oncogenesis.

#### 1. Meta-Analytic Cognitive Activation Maps

The Neurosynth Cognitive Atlas was harnessed to obtain probabilistic measures of association between voxels and higher-order cognitive processes^36^. Neurosynth is a meta-analytic platform that collates results from over 15, 000 peer-reviewed and published functional MRI studies by searching key words – these maps are published alongside functional MRI voxel coordinates (github.com/neurosynth/neurosynth). The output from this database is the probability that a specific cognitive process if there is observed activation at a given voxel. While there are several hundred cognitive processes accessible on Neurosynth, we focused on three specific terms that are conventionally associated higher-order executive function^2^: 1) working memory, 2) inhibitory control, and 3) cognitive flexibility. The extracted coordinates for each cognitive processes from Neurosynth were then parcellated based on the Glasser-MMP HCP atlas and z-scored^37^. These meta-analytic activation maps derived from Neurosynth were interpreted as brain regions functionally linked to the cognitive task.

#### 2. Normative Diffuse Glioma Tumour Frequency Map

Data from low grade gliomas (LGGs) were accessed from the Multimodal Brain Tumour Image Segmentation Challenge 2019 database^38–40^. T1-weighted images from 101 low-grade gliomas (age=46.7, male=45) were segmented by board-certified radiologists, which denoted voxels that were contrast enhancing tumour, non-enhancing core, and oedema. Scans were collected at 19 different sites with different acquisition parameters; however, the structural imaging were processed through the same pipeline and undergoing registration to a common template with resample at 1mm^3^ isotropic resolution. Tumour masks were combined across the entire cohort to create a group-averaged tumour frequency map in the same space. This pre-processed data was accessed on the publicly available BRaTs tumour dataset website.

#### 3. Neurobiology of the MD System

To understand the neurobiology underpinning the multiple demand (MD) system and how this network can simultaneously support higher-order cognition and gliomagenesis, we leveraged Cam-CAN dataset to map functional hubs, the Allan Human Brain Atlas (AHBA) to map gene expression data^41^, neurotransmitter data from Positron Emission Tomography (PET) data scans, and normative biological maps from the Penn Lifespan Informatics and Neuroimaging Center (PennLINC) on aerobic glycolysis, cerebral blood flow, allometric scaling, evolutionary hierarchy, and externopyramidization^56,70^.

##### Functional Hubs

Since the relationship between functional connector hubs and their spatial localization relative to the MD core remains unclear, the Cam-CAN dataset was used generate a group-averaged functional connector hub map. This map was constructed by averaging connectivity matrices across the lifespan and applying the Louvain algorithm for community detection. Considering the MD system’s role in domain-general cognitive tasks and its widespread distribution across the cortical surface, we hypothesized that MD nodes would more likely serve as connector hubs than non-MD hubs.

##### Cell-Type Specificity

To investigate the cell-type specificity associated with the MD system, we utilized the Allen Human Brain Atlas (AHBA), which provides transcriptomic bulk-RNA sequencing microarray data. The Abagen toolbox and pipeline were employed to process the microarray data^43^, which was subsequently parcellated using the MMP 1.0 mm Glasser parcellation scheme. Using Seliditz et al.’s compilation of gene sets for various cell types—including oligodendrocytes, oligodendrocyte precursor cells (OPCs)^44^—we systematically assessed whether the MD core showed preferential enrichment in specific cell types. Given the growing body of evidence from animal and human studies highlighting myelin plasticity as a critical regulator of cognitive functioning^45–47^, we hypothesized that oligodendrocytes and OPCs would be uniquely enriched in the MD core network but not in other networks, such as the CON and DMN.

##### Gene Ontology

Using the AHBA microarray expression data comprising 20,467 genes across 20 MD core brain regions^48^, we conducted a K-means clustering analysis to identify gene clusters expressed within the MD system. The resulting clusters were then analyzed using the GOrilla gene ontology database to identify the physiological processes associated with regulating the MD system^49^.

##### Neurotransmitter Density

Building on decades of evidence linking dopaminergic and serotonergic neurotransmission to both improvements and deficits in executive functioning^50–55^, we utilized publicly available positron emission tomography (PET) maps to examine their co-localization with the MD system^30^. PET imaging uniquely enables the estimation of in vivo neurotransmitter receptor concentrations across the entire brain, a capability not offered by other imaging modalities. Glutamatergic data served as a control in this analysis. Specifically, we used recently released and publicly available PET tracer data to evaluate the relative spatial distribution of these neurotransmitters^42^.

##### Diverse Biological Maps

The Penn Lifespan Informatics and Neuroimaging Center (LINC) recently released maps highlighting various neurobiological properties^56^. These maps detail the regional spatial distribution of aerobic glycolysis, cerebral blood flow, allometric scaling, evolutionary expansion, and the first principal component of functional connectivity. We analyzed these maps to assess their co-localization with the MD system and its subnetworks. Given the extensive and highly reproducible evidence showing that the MD system is consistently activated across a wide range of cognitive tasks, we hypothesized that aerobic glycolysis and cerebral blood flow would be highest in MD core regions. Additionally, since higher-order cognition distinguishes humans from non-human primates and lower-order mammals (e.g., rodents)^57^, we expected the most evolutionarily expanded cortical regions to align closely with the MD system. Finally, based on Mesulam’s hierarchical model and the idea that higher-order cognitive regions occupy the top of this hierarchy^58,59^, we hypothesized that the first principal component of functional connectivity would co-localize with the MD system, complementing our earlier findings on cortical manifolds^58^.

##### Glioma Proto-Oncogenes

we examined whether 14 glioma proto-oncogenes co-localized with the MD core system using the spin test described below. Gene expression maps were obtained from the Allen Human Brain Atlas and averaged. The panel of genes analyzed included TP53, NF1, MDM2, PIK3CA, FUBP1, NOTCH1, PDGFRA1, RIS, PI3K, IDH1, IDH2, TERT, ATRX, EGFR, CDKN2A, CDKN2B, PTEN, and RIK—genes known to be amplified, deleted, or mutated in specific types of gliomas^60^.

#### 4. Spatial Permutation Testing

Spin tests, also known as spatial permutation tests. are used to assess the spatial similarity between two maps while preserving the covariance structure^61^. Spin tests have become a widely used analytical tool in systems neuroscience for statistically comparing different brain ^42–44^. However, this study marks the first time this approach has been applied to investigate the MD system, particularly in the context of the most recent fine-grained MD parcellation^23^. For further discussion on null models in network neuroscience, see reference 62.

To evaluate the hypotheses outlined earlier, we utilised the spin test developed by Alexander-Bloch and colleagues. This method randomizes the relationship between the network (i.e. brain topology) and annotations (i.e, gene expression data) by randomly rotating spherical projections (or spin) of brain annotation maps. Each coordinate is then matched to its nearest rotated counterpart, resulting in a randomized correspondence between network nodes and annotations^62^. In these analyses, a modification was made for comparing a binary (MD system) and a continuous map. Instead of correlating the real data with a null (spun) distribution of correlation coefficients, we compared the real sum of z-scores against a null (spun) sum z-scores. The null distribution was built by rotating the binary map and repeating this procedure 10,000 times to determine if the empirical sum of z-scores is in the top 5% of this distribution. Given that correlation coefficients rely on rank and are unsuitable for binary maps like the MD system, we reported the sum of z-scores instead. P-values were corrected for multiple comparisons via the Benajmin-Hochberg method.

### Section B Linking Hub Architecture with Cognition, Gliomas, and Surgical Intervention

Given the inherent difficulties of mapping the MD system longitudinally in neurosurgical patients and the close relationship between task-evoked and task-free brain functional regions, we first sought understand the relationship between the brain’s MD system and mesoscale hub architecture in healthy individuals across the lifespan (Figure 1A). We then sought to understand how glioma infiltration of the MD system affected cognitive outcomes and hub plasticity (Figure 1B). Using these insights, we then leveraged a rare neurosurgical cohort undergoing neurosurgery to determine how diffuse gliomas integrate within the brain’s hub architecture and how neurosurgical resection of gliomas affect hub topology and long-term cognitive outcomes.

#### 1. Healthy Lifespan Cohort (Cam-CAN) and Longitudinal Neurosurgical Cohort (CAESAR)

##### Cambridge Centre for Ageing and Neuroscience (Cam-CAN) Cohort

Data from healthy participants across the lifespan were acquired as part of the Cambridge Centre for Aging and Neuroscience (Cam-CAN) dataset which comprised of healthy individuals aged 18-88 years. The study has been described in detail elsewhere^63^. Cam-CAN (n=620) participants and our surgical cohort (described below) were scanned on the same Siemens Prisma 3T MRI scanner with the same acquisition parameters. Thus, Cam-CAN participants were propensity-score matched to our surgical cohort in a 1:10 ratio for age (+/- 1) and sex and formed the control sample. The Cattell fluid intelligence (total) score, which measures executive functioning and the ability to solve novel reasoning problems^34^, was utilized as the dependent variable when link Cam-Can data to neuroimaging data.

##### CAESAR Experimental Medicine Study

In the CAESAR prospective, longitudinal, experimental medicine study of patients undergoing diffuse glioma surgery, participants were recruited by the adult neuro-oncology multidisciplinary team at Addenbrooke’s hospital (Cambridge, UK). The study was approved by the Cambridge Central Research Ethics Committee (Reference number 16/EE/0151) and all patients provided written informed consent. Participant characteristics, inclusion and exclusion criteria, and image scanning parameters have been previously described^64^. Connectomic data was acquired pre-surgery, postoperative Day 1, and 3 and 12 months in the rehabilitation period; four time points.

#### 2. MRI Pre-Processing, Tumour Mask and Overlapping Calculation, and Functional Time Series Extraction

Details of structural MRI processing, lesion marking, resting-state functional MRI (rsfMRI) pre-processing, and lesion masking have been previously reported^64^. Importantly, the Glasser parcellation scheme from the Human Connectome Project (HCP) was chosen to parcellate the cortex into 360 defined regions to establish a common nomenclature with other neurosurgical connectomic studies^65–67^.

In brief, functional MRI (fMRI) data from patients with diffuse gliomas were collected at the Wolfson Brain Imaging Centre, University of Cambridge. Scans were performed using a Siemens Magnetom Prisma-fit 3 Tesla machine equipped with a 16-channel receive-only head coil (Siemens AG, Erlangen, Germany). Single-echo fMRI sequences were selected to facilitate clinical translation and simplify preprocessing. During the same session as structural MRI (sMRI) acquisition, resting-state fMRI (rsfMRI) data were obtained while patients kept their eyes closed. The sequence was BOLD-sensitive with parameters: TR = 1060 ms, TE = 30 ms, acceleration factor = 4, flip angle = 74°, voxel size = 2 mm³, field of view = 192 × 192 mm², and acquisition time = 9 minutes and 10 seconds. Preprocessing of fMRI data was performed using FSL’s MELODIC independent component analysis (ICA). Noise components identified during this process were removed using ICA-FIX, trained specifically for this dataset. Additional standard preprocessing steps included slice-timing correction, bias field correction, rigid body motion correction, image normalization by a single scaling factor, and spatial smoothing with a 5 mm full-width at half-maximum kernel to account for residual variability. Subsequent analyses focused on physiologically relevant BOLD signal frequencies (0.03–0.12 Hz) using wavelet filtering (wavelet scales 3 and 4).

The fMRI data were linearly registered to T1-weighted structural images using six degrees of freedom with the Advanced Normalization Tools (ANTs) platform. The resulting transformations were used to inverse-map the Glasser parcellation scheme into the fMRI space, allowing the extraction of average time series for each parcel. To address head motion artifacts, framewise displacement (FD) was calculated, with frames exceeding FD > 0.4 identified as outliers. The frame preceding and two frames following each outlier were also excluded due to delayed motion effects on the BOLD signal. Scans with more than 50% outliers were excluded from analyses.

Across 17 patients scanned at four time points (pre-surgery, post-surgery Day 1, post-surgery Month 3, and post-surgery Month 12), a total of 55 usable scans (∼85%) were included. Exclusions resulted from missed follow-up appointments, logistical issues, or excessive head motion. Longitudinal data were available for the following time points: pre-surgery (n = 17), post-surgery Day 1 (n = 17), post-surgery Month 3 (n = 13), and post-surgery Month 12 (n = 11). Ultimately, 11 patients had complete neuroimaging datasets across all time points.

#### 3. Modularity, Connector Hubs, and Graph Theoretical Analyses

Following the preprocessing of the rsfMRI data, we exclusively used the resulting 360×360 weighted functional connectivity matrices, with all negative elements set to zero. Graph theory analyses were conducted using the Brain Connectivity Toolbox (BCT)^40^. To assess the inter- and intra-modular degree of each network node, we first determined the network’s community structure by performing a consensus modular decomposition. This involved 1,000 iterations of the Louvain clustering algorithm, with the resolution parameter (γ) set to 1. The Louvain algorithm produced two key outputs: Newman’s modularity score (Q) and the community affiliation vector (Ci). Newman’s Q, calculated as shown in Equation 1, quantifies the global modularity of the network by measuring the strength of the division into modules. Networks with high modularity exhibit dense intra-module connections and sparse inter-module connections. Specifically, In specific, Aij is the adjancey matrix with elements representing weights between I and j. Pij is the probability that nodes i and j are connected in a random null network, γ is the resolution parameter, ci is the community affiliation vector of each node.

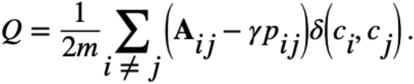

At the nodal level, utilising the community affiliation (Ci) vector from the Louvain algorithm, we estimated the participation coefficient (Pc) which is widely used as a measure of modular community structure (Equation 1)., where *k_i_(m*) is the number of edges between node *i* and all nodes in module *m*, and the sum is overall M modules. If a node’s Pc is close to zero, then it is mostly connected to other nodes in the same module and lacks significant connections to other modules. While if a node’s Pc is close to 1, then it is considered to be significantly connected to all modules throughout the global network. An intuitive example would be Heathrow airport has a higher Pc in the global airport network given the long and short-haul flight connections, whereas Stanstead airport would have a significantly lower Pc given it’s more domestic and short-haul flight connections.

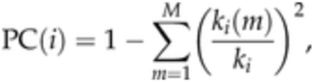

Connector hubs were defined as the top 20th percentile of nodes ranked by Pc, consistent with the “diverse club” definition frequently used in prior studies. Notably, baseline Q and Pc metrics have been shown to predict cognitive plasticity following interventions across various neurological conditions. Therefore, we focused our analyses on these clinically significant graph-theoretical metrics.

#### 4. CAESAR Neuropsychological Assessment and Linkage to Neuroimaging Data

Neuropsychological assessment was conducted utilizing a battery of tests that assessed multiple cognitive domains, including verbal memory, verbal skills, nonverbal memory, attention, and executive function. The details of each test are described in the Supplementary Material (Table S1). Patients underwent cognitive assessments two weeks prior to surgery and at 3-month and 12-month intervals following surgery. A cognitive deficit was defined, as in earlier research,^39,40^ as a performance score at least two standard deviations below the mean of a reference population for any given test. The total number of deficits was calculated as the sum of tests where a patient’s score fell below this threshold. The number of acquired cognitive deficits was determined by subtracting the total number of presurgical deficits from the total number of postsurgical deficits. A positive value indicated cognitive deterioration, while a negative value indicated cognitive recovery. This composite score, spanning all domains, served as a measure of overall higher-order cognition and dependent variable for subsequent neuroimaging analyses.

#### 5. The Effect of Glioma Invasion of the MD System and Hub Architecture

Given the MD system’s seminal role in higher-order cognition, we evaluated how glioma infiltration impacted cognitive function. Specifically, for each CAESAR patient, the percentage overlap of their glioma with the MD system was quantified. Subsequently, the change in cognitive function was calculated for each patient as the difference between the number of cognitive deficits at 12 months post-surgery and the number recorded pre-operatively. This data was then used to conduct a binomial logistic regression analysis, examining the relationship between MD core system overlap and long-term changes in cognitive function, either improvement or decline. Additional sensitivity analyses using the MD penumbra system and non-MD regions.

To assess the impact of gliomas on global modularity (Q), we compared each patient’s pre-surgical Q to that of age- and sex-matched healthy controls. Non-parametric t-tests were used for these comparisons. Additionally, using an independent Belgian dataset^66^, we replicated this analysis and included comparisons with meningioma patients. For a more detailed analysis, we examined hub topology differences between patients and healthy controls. Spatial autocorrelation testing was employed to identify disruptions in hub patterns between the groups. Lastly, we analyzed how intratumoural presurgical participation coefficient differed in patients with long-term improvements and deficits.

#### 6. The Effect of Resecting Connector Hubs on Modularity and Long-Term Cognitive Outcomes

To assess the presurgical utility of rsfMRI to ideally complement DES, we exclusively analysed presurgical functional imaging data to make long-term cognitive predictions. Specifically, we assessed the impact of resecting connector hubs in the intratumoural and peritumoural areas and how this affected long-term cognitive outcomes (defined as change from presurgical and postsurgical cognitive functioning). Patients with long-term cognitive improvements were defined as having zero or fewer number of deficits compared to pre-surgery while patients with long-term cognitive deficits were defined as having new or increased number of deficits. The analyses of connector hub resection were contrasted with resecting provincial hubs.

Based on the comprehensive review by Gallen and D’Esposito^33^ , we tested the hypothesis that ‘intervention-related’ modularity can predict long-term cognitive outcomes. To this end, we compared baseline contrahemispheric modularity in the perioperative period to determine if this can serve as a biomarker for patients who may benefit from interventional neurorehabilitation in the perioperative period^67^. We focused on the contrahemispheric hemisphere order to avoid the effects of the tumour, mass effect, and oedema in the ipsilateral hemisphere and for the goal of facilitating contralateral interventional neurorehabilitation in future studies. To achieve, we split our patient cohort into two: long-term cognitive improvers and decliners. Individual and summary-level data of each patient is tabulated (Table S1 and Table 2). From here, we worked backwards to determine from the preoperative stage, how contrahemispheric modularity (Q) changed in both cohorts over time. Linear Mixed Effects modelling was utilised to assess any significant differences over time.

## Results

### Meta-Analytic Cognitive Activation Maps

Using Neurosynth, a meta-analytic tool for functional neuroimaging data, a set of frontal and parietal brain regions are consistently activated co-activated across three different cognitive tasks: attention, working memory, and cognitive control (Figure 2). These terms were chosen given they have been operationalized from the seminal work from by Miyake and Friedman^2^. Spin test between each map and the MD system revealed a significant positive correlation between the MD core and cognitive action maps for attention (r=0.623, p=0.023), working memory (r=0.742, p=0.013), and cognitive control (r=0.562, p=0.015). All p-values were corrected for multiple comparisons via benjamin-hochberg method.

**Figure 2:**
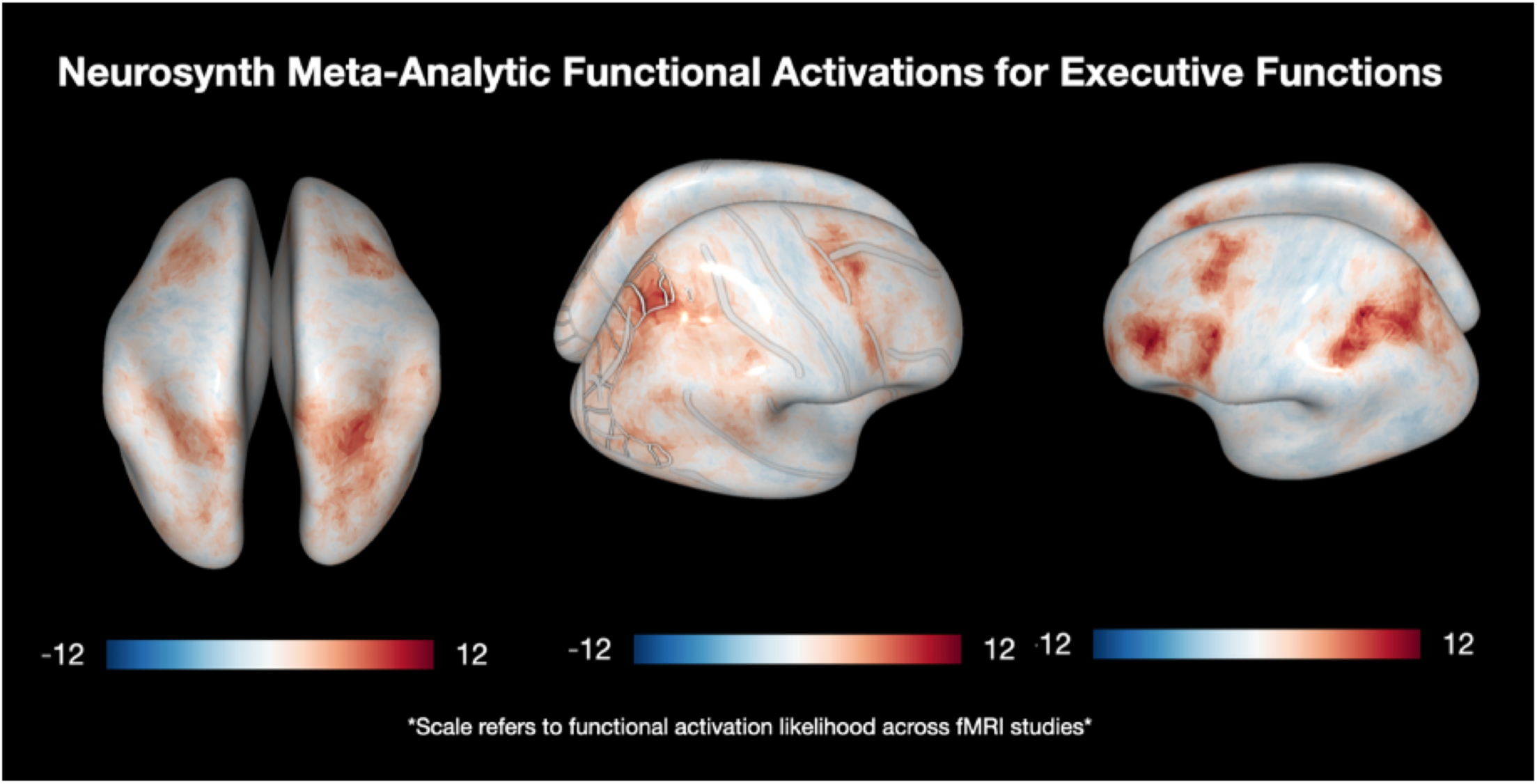
Using the neurosynth meta-analytic platform, search terms for attention, working memory, and inhibitory control are present left to right. Based on ∼1200 task-based functional MRI studies, a distributed frontoparietal network consistently emerged (highlighted as darker red areas). Spin tests results: attention (r=0.623, p=0.023), working memory (r=0.742, p=0.013), and working memory (r=0.772, p=0.015). All p-values corrected for multiple comparisons via benjamin-hochberg method.

### Glioma Vulnerability Map and Proto-Oncogene Expression

The tumour frequency map (Figure 3-middle) of low-grade gliomas demonstrated a strong spatial correlation with the MD core system (z = 0.832, p = 0.042) and a weaker correlation with the MD penumbra (z = 0.453, p = 0.0012). In contrast, analyses conducted with non-MD regions showed no significant spatial correlation (z = 0.223, p = 0.241). Furthermore, the MD core system was significantly correlated with mean proto-oncogene expression (z = 0.622, p = 0.032), whereas no significant correlation was observed for the MD penumbra (z = 0.132, p = 0.192) or non-MD regions (z = 0.232, p = 0.32). All p-values were corrected via the Benjamin-Hochberg method. Notably, the glioma patient cohort from the BRaTS dataset included individuals treated with both operative and non-operative approaches, minimizing the likelihood of recruitment bias in these findings.

**Figure 3:**
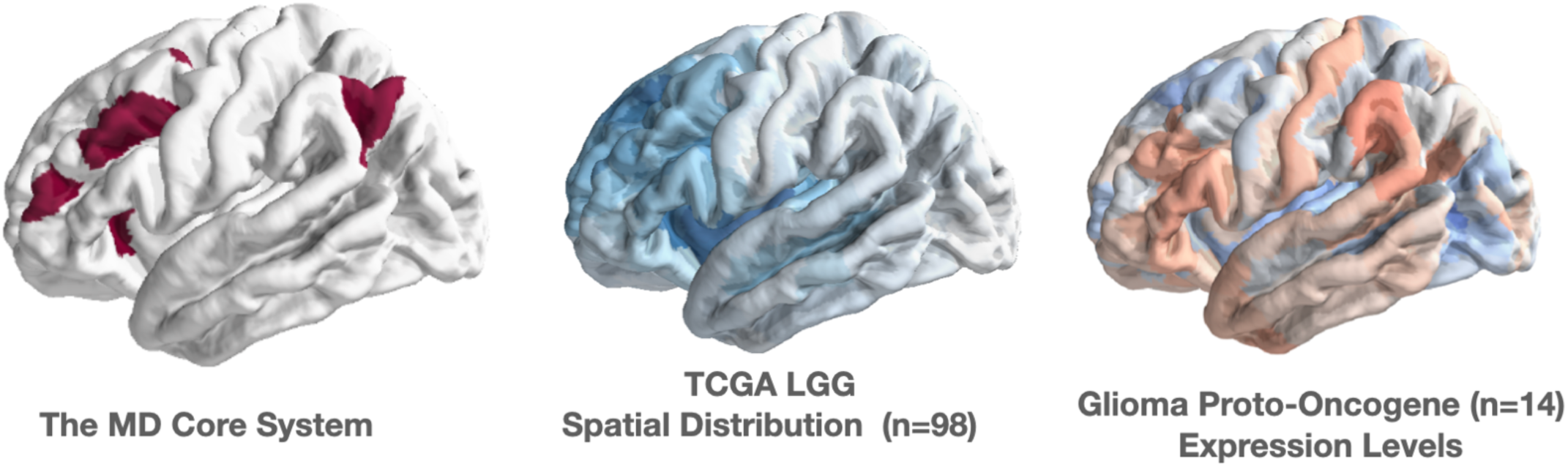
Left to Right. A) Canonical MD Core System as mapped Assem and colleagues. MD Penumbra not illustrated here. B) Glioma distribution map acquired from The Cancer Genome Atlas (TCGA). C) Mean gene expression map of 14 proto-oncogenes involved in gliomagenesis. Spin test reveals gliomas preferentially infiltrate the MD Core system, rather than the MD Penumbra or non-MD brain regions. In addition, glioma proto-oncogenes are preferentially expressed in the MD Core, rather than the MD penumbra or non-MD brain regions. All p-values corrected for multiple comparisons via benjamin-hochberg method.

### Neurobiology of the Multiple Demand Network

First, the MD core system showed a significant spatial correlation with functional connector hubs derived from group-aggregated rsfMRI data (z = 0.93, p = 0.032), whereas the MD penumbra was comparably (z = 0.57, p = 0.02). Second, oligodendrocyte precursor cells (OPCs), but not oligodendrocytes, were uniquely enriched in the MD core (z = 0.4855, p = 0.0117). Interestingly, the MD CON also exhibited OPC enrichment (z = 0.72455, p = 0.0089), while the MD DMN was significantly enriched with oligodendrocytes (z = 1.0309, p = 0.04) (Figure 4B). Third, allometric scaling, cerebral blood flow, evolutionary expansion, the principal gradient of functional connectivity, and aerobic glycolysis were all significantly co-localized with the MD core (see Supplementary Material for z-scores and adjusted p-values). Additionally, serotonergic 5HT1b receptors were highly enriched in both the MD core and MD CON (Figure 4B). Finally, K-means clustering of genes differentially expressed in the MD core revealed two distinct clusters (Figure 4C). Gene ontology analysis indicated that Cluster 1 was associated with regulatory and metabolic processes, particularly WNT signaling (p = 2.84e-35), while Cluster 2 was linked to neuropeptide signaling (p = 1.09e-33). The networks MD core, MD CON, and MD DMN were selected based on their well-documented and distinct functions in the literature, with the latter two serving as controls for comparisons with the MD core.

**Figure 4.**
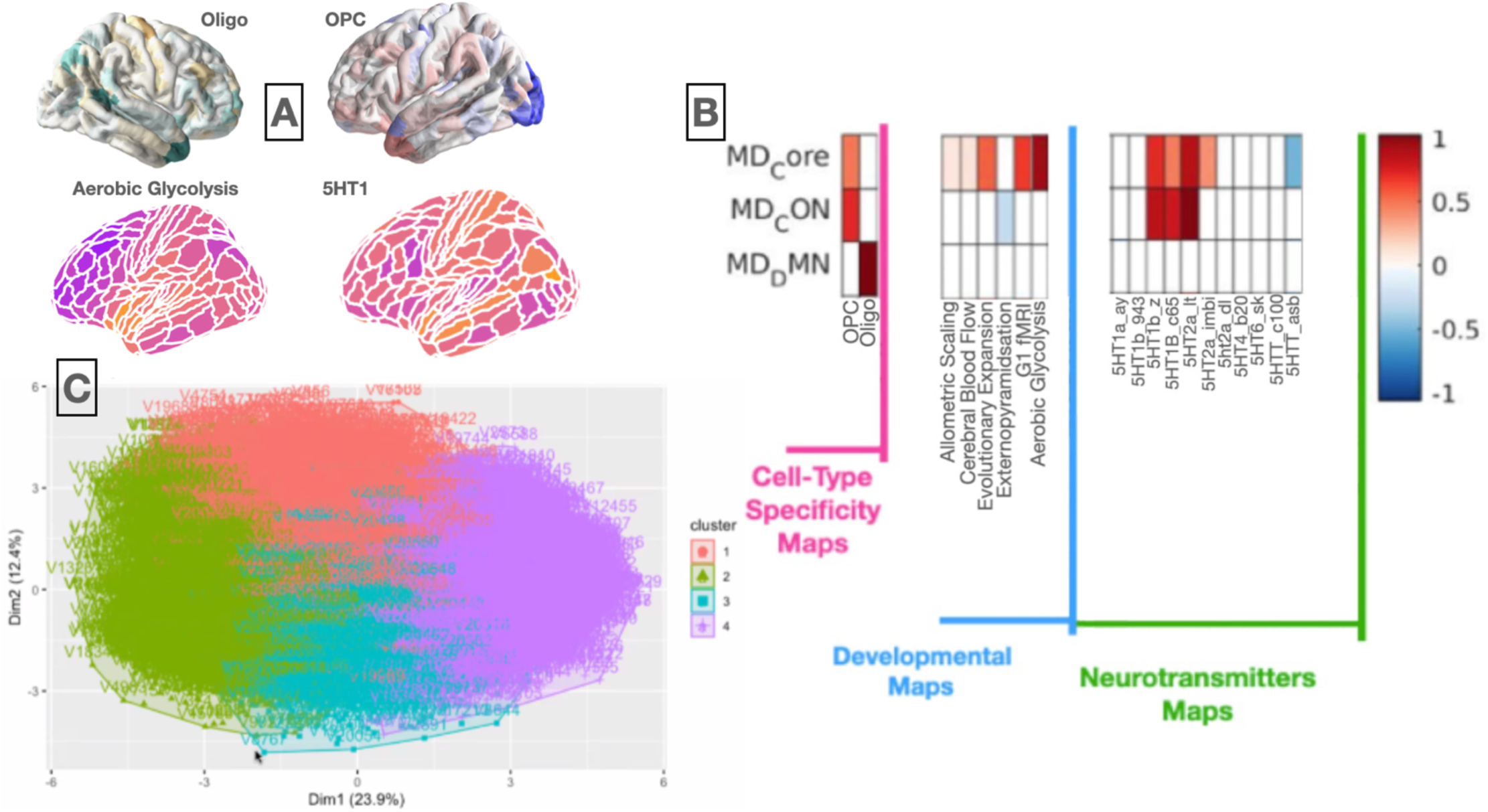
Neurobiological Characterisation of the MD system. A) Cortical maps a few biological features (oligodendrocyte cells, OPCs, aerobic glycolysis, and 5HT1 gene expression). B) Results summary of the spin tests between networks and biological features. Only significant p-values after adjustment displayed; axis refers to the strength of the relationship. C) The output of the k-means clustering algorithm revealed 4 distinct gene sets involved in the MD core system. Gene ontology analyses revealed Cluster 1 (red) representing regulatory and metabolic processes and Cluster 2 (green) representing genes involved in neuropeptide signalling.

### Frequency of Connector Hubs in the MD System

In the CamCAN cohort (n = 629, mean age = 54.6, age range = 18–88, female = 51%), multiple linear regression controlling for age, sex, and age*sex interactions revealed a negative association between modularity (Q) and executive functioning, as measured by the Cattell Fluid Intelligence Test (Figure 5A) (R² = 0.453, p = 0.0013 for Q covariate, slope = -8.790). This relationship showed only a slight improvement when using a generalized additive model (R² = 0.483) to account for both linear and non-linear effects of age on modularity.

**Figure 5.**
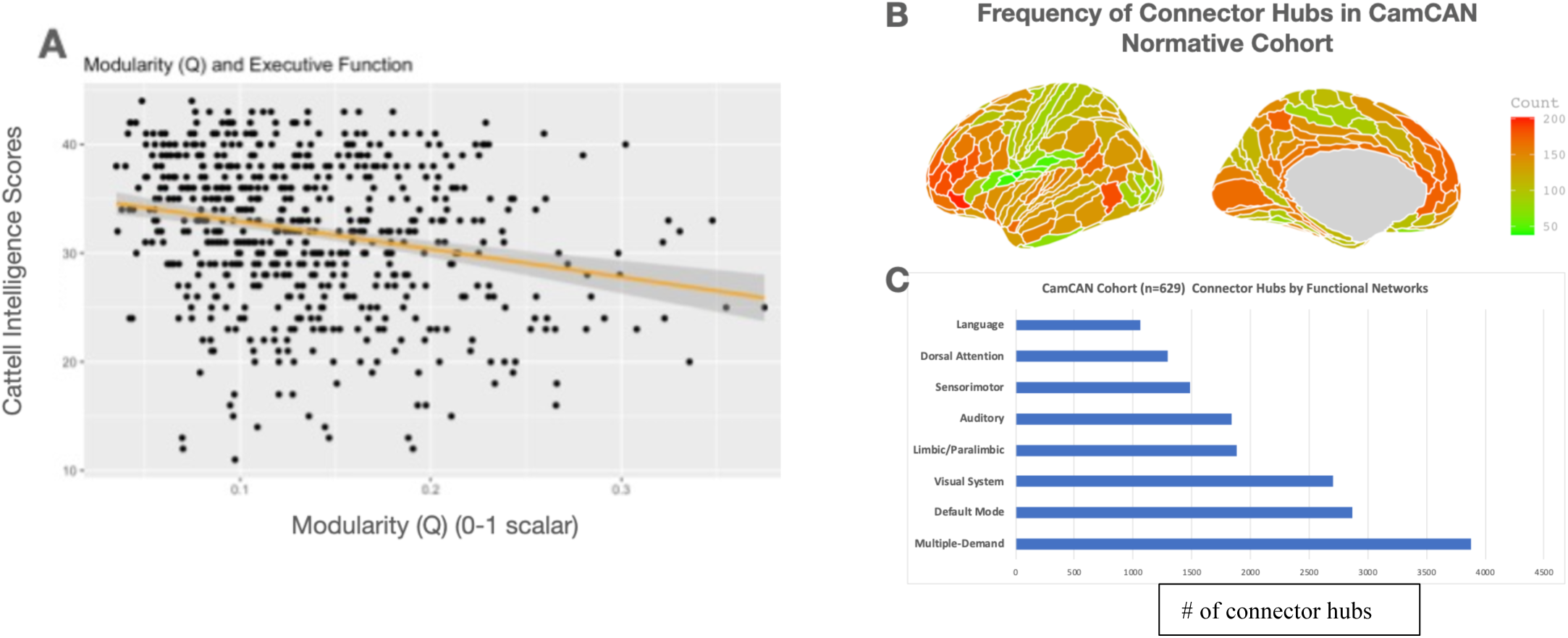
The Modular Organisation of the Brain Predicts Executive Functioning in Health Individuals Across the Lifespan. A) Modularity is negatively associated with executive functioning in healthy individuals across the lifespan, after controlling for age and sex. B) The prefrontal and parietal regions exhibited the highest frequency of connector hubs across the entire cohort. C) The MD system had the greatest number of connector hubs, followed by the default mode network.

When examining the location of connector hubs across the cohort, the majority were found in the medial and lateral prefrontal cortex and parietal regions, highlighting brain areas most diversely integrated with other modules (Figure 5B). Analysis of the network identity of these connector hubs revealed that most belonged to the Multiple Demand (MD) system, followed by the default mode network and the visual system (Figure 5C). Notably, individuals with a greater number of connector hubs in the MD system tended to exhibit higher executive functioning, aligning with prior research on task-positive and task-negative functional neuroimaging and their relationship to cognitive performance.

### The Effect of Glioma Invasion on the MD System

To assess the cognitive impact of gliomas infiltrating the MD core, penumbra and system, a Spearman correlation was used to analyze the relationship between tumour overlap and changes in long-term cognitive outcomes. Interestingly, greater pre-surgical glioma infiltration in the MD core system was associated with fewer long-term cognitive deficits (Spearman’s rho = -0.75, p = 0.0005) (Figure 6A). A similar but weaker association was observed with tumour infiltration into the MD penumbra (rho = -0.57, p = 0.01) (Figure 6B). In contrast, tumour overlap in non-MD regions showed no significant relationship with long-term cognitive outcomes (Spearman’s rho = -0.11, p = 0.3) (Figure 6C). P-values were corrected for multiple comparisons via the Benjamin-Hochberg methods. These findings suggest that the canonical MD system may undergo functional plasticity to accommodate glioma infiltration, thereby minimizing disruptions to long-term cognitive functioning. Table S1 summarizes the long-term behavioural outcomes of the CAESAR cohort.

**Figure 6.**
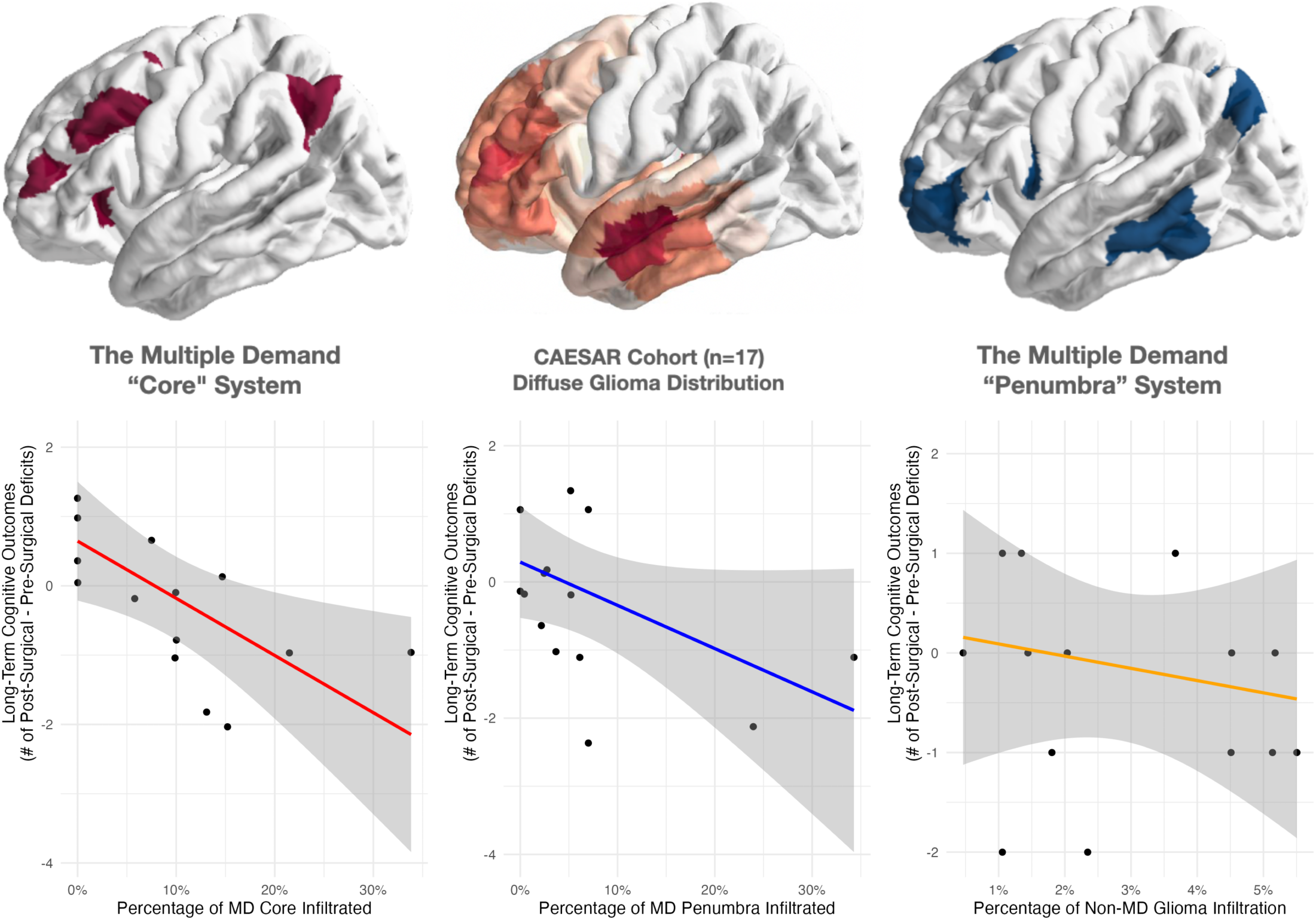
Glioma Invasion into the MD System Top Panel: *From Left to Right: Canonical MD Core system, CAESAR Cohort’s glioma spatial distribution,and canonical MD Penumbra system. **Bottom Panel*** A) Relationship between the percentage of presurgical tumour infiltration in the MD Core system (x-axis) and the number of long-term cognitive deficits (y-axis), calculated as the difference between post-surgical and presurgical deficits. The two variables are strongly negatively correlated (rho = -0.75, p = 0.0005). B) Relationship between the percentage of presurgical tumour infiltration in the MD Penumbra system (x-axis) versus the number of long-term cognitive deficits (y-axis). A moderate negative correlation is observed (rho = -0.57, p = 0.01). C) This graph depicts the percentage of presurgical tumour infiltration in non-MD system regions (x-axis) against the number of long-term cognitive deficits (y-axis). The correlation is weak and statistically insignificant (rho = -0.11, p = 0.3).

### Modularity in Glioma Patients over time

To better understand how increased glioma infiltration of the MD system leads to decreased long-term cognitive deficits, we assessed global modularity in patients by comparing them to age- and sex-matched controls. There was no significant group difference between patients and controls, as assessed by the Wilcoxon unpaired t-test (p=0.891) (Figure 6A), suggesting that the patients’ brains are reorganising at the nodal level to compensate for the presence of glioma and maintain modular architecture. Moreover, when assessing whole-brain modularity over time, on-way ANOVA revealed no significant group differences (p=0.642). Lastly, there was a significant difference in presurgical intra-tumoural participation coefficient in patients with long-term improvement and decline (p=0.016) (Figure 7B).

**Figure 7.**
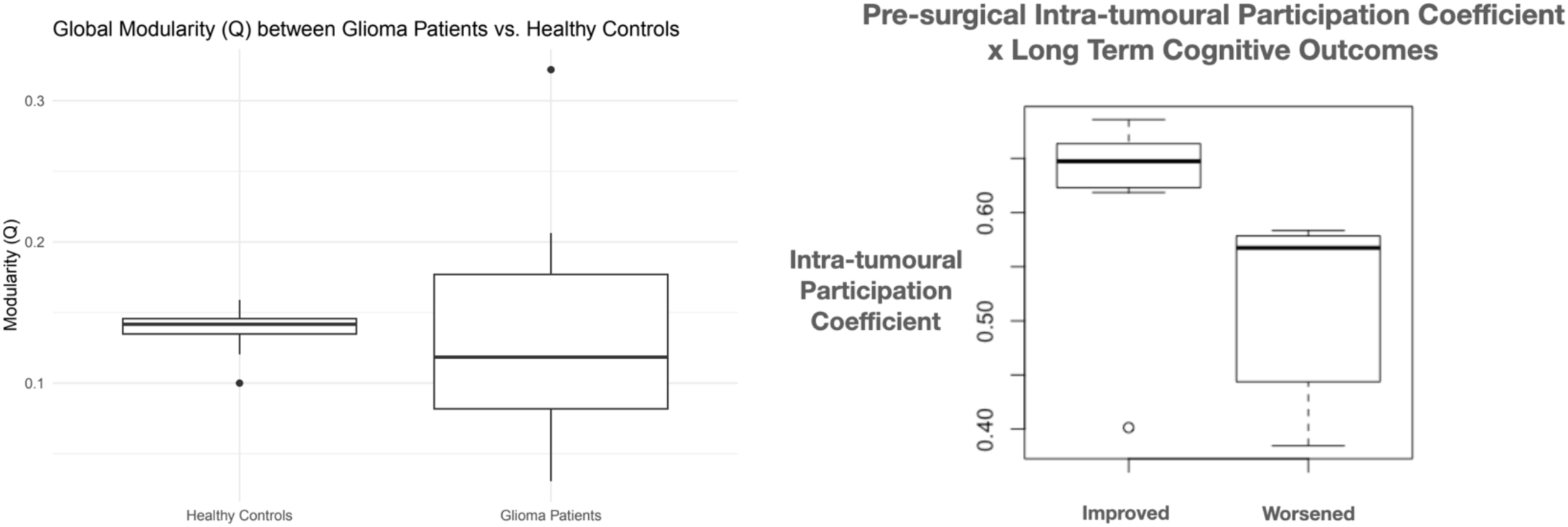
The Effect of Cancer on Global Modularity. A) Global modularity compared between age- and sex-matched healthy controls (mean=0.142, SD=0.01) and glioma patients (mean=0.134, SD=0.073). B) Significant difference when comparing presurgical intratumoural participation coefficient in patients with long-term improvements and decline (p=0.016)

### The Effect of Resecting Connector Hubs on Modularity and Long-Term Cognitive Outcomes

Since connector hubs are critical for maintaining the modular organization of a network, understanding the impact of their removal is essential for surgical planning. Resecting more than five connector hubs was associated with long-term cognitive deficits, whereas removing four or fewer resulted in long-term cognitive improvements (Figure 8A, p = 0.0211). In contrast, resecting varying numbers of provincial hubs had no significant effect on long-term cognitive outcomes (Figure 8A, p = 0.344). Notably, there was no significant difference in tumour volumes between the two groups (p = 0.5303).

**Figure 8.**
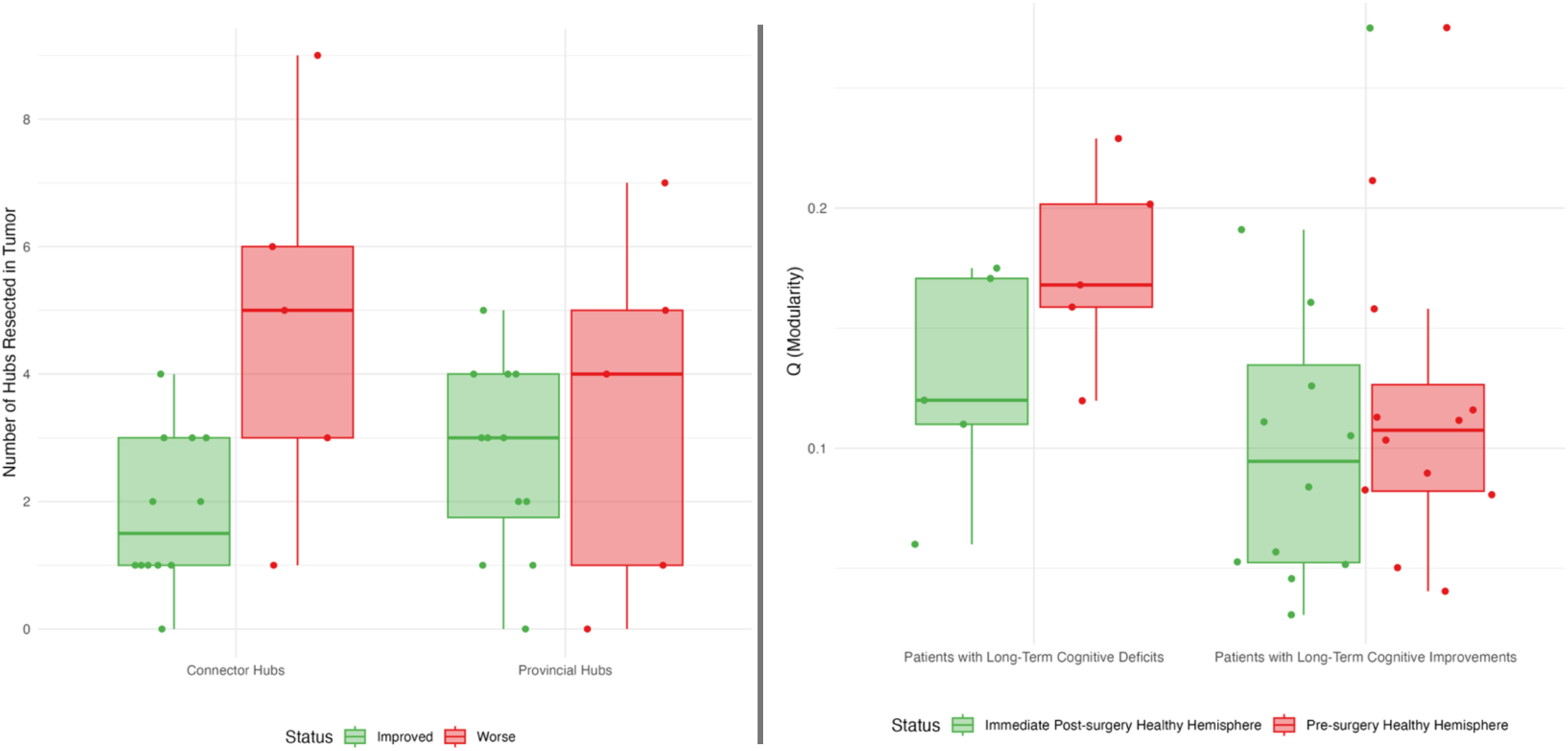
Left Panel-Impact of Resecting Connector Hubs on Long-Term Cognitive Outcomes. A) Patients with long-term cognitive decline (n = 5) had significantly more connector hubs resected during surgery compared to those with long-term cognitive improvements (n = 12) (Cohen’s d = 1.3, p = 0.0211). B) In contrast, assessing the number of provincial hubs resected revealed no significant differences between the two groups (p = 0.344). **Right Panel-Longitudinal Modular Network Trajectories Based on Cognitive Outcomes.** B) Patients who experienced long-term cognitive deficits after surgery showed a significant decrease in modularity (Q) immediately post-surgery (Cohen’s d = 0.76, p = 0.031). B) In contrast, patients with long-term cognitive improvements did not exhibit a decrease in modularity (Q) immediately after surgery.

To evaluate whether global modularity could serve as a biomarker for long-term cognitive outcomes, we analyzed Q in the contralateral hemisphere of patients who showed long-term cognitive improvement and those who did not. Patients with persistent cognitive deficits (n=5) exhibited a significant decrease in perioperative modularity (Q) (p=0.031), whereas patients with cognitive improvements (n=12) showed no significant change in perioperative modularity (p=0.631). Statistical analysis was performed using non-parametric Mann-Whitney U tests.

Importantly, in patients stratified as with long-term improvements or long-term decline, there was no significant difference in pre-surgical tumour volume (p=0.2468), IDH1 mutation status (p= 0.3827), and medical treatment decisions (i.e. chemotherapy or observation, p=0.1763) as tested by non-parametric t-tests.

## Discussion

In a series of interconnected investigations, we aimed to develop a cellular to circuit level explanation to how the MD system supports both higher-order cognition and gliomagenesis. We first found that brain regions commonly plagued by gliomas spatially correlate with MD core network responsible for domain-general cognition. Using the Allen Human Brain Atlas (AHBA), we showed that glioma proto-oncogenes and OPCs are preferentially expressed in the MD system compared to other canonical brain networks (i.e, default mode network). Furthermore, we demonstrated that metabolically expensive connector hubs are a key feature of the MD system and that glioma infiltration into this network causes minimal cognitive deficits by preserving the modular organisation of the brain. Finally, surgical resection of connector hubs leads to long-term cognitive decline and perioperative modularity may serve as a biomarker for long-term cognitive outcomes.

### The Glioma & MD System Interface

The observation that gliomas spatially co-localise with the MD system is consistent with recent studies demonstrating that glioma frequency is elevated in the association cortex and correlates with graph-theoretical metrics for highly connected brain areas ^11,43^. The work presented in this the study replicates and builds upon existing knowledge work by employing a more granular parcellation scheme (the Glasser HCP) while employing an anatomically-defined approach to the MD system. Additionally, we found that glioma proto-oncogenes and OPCs are enriched in the MD core system, rather than the MD penumbra, suggesting that these regions are potentially more vulnerable to gliomagenesis. Overall, this supports the hypothesis that highly active brain hubs, which are metabolically demanding, are particularly susceptible to not only neurological conditions (i.e., stroke) and psychiatric illnesses (e.g., schizophrenia)^71^, but also vulnerability to gliomagenesis.

When evaluating the effects of glioma infiltration (n=17) in the MD system, greater infiltration was associated with better long-term cognitive outcomes. This outcome appears to be linked to the rearrangement of connector hubs, which helps preserve the brain’s modular architecture while adapting to the presence of the glioma. Since gliomas require significant energy and metabolic resources to grow, it is possible that the connectome compensates by redistributing some of its energy to the tumour. It is interesting that the greater variance observed in the presurgical modularity (Q) of glioma patients compared to age- and sex-matched healthy controls; perhaps there is individualised approach to hub-rearrangement based on the tumour’s molecular, histological, and spatial location. Further studies with larger sample sizes are needed to clarify the mechanisms of presurgical compensation and their impact on long-term outcomes.

### Can thought itself drive glioma growth?^72^

The importance of activity-dependent myelination on learning has recently taken a dominant perspective in neuroscience. Specifically, OPCs have been demonstrates to play a key role in myelin plasticity, promoting the proliferation of OPCs, generating new oligodendrocytes, and shaping myelin during development and learning. This is against the backdrop that neuronal-glial interactions have been demonstrated to shape circuit dynamics and pruning given that glial cells can also form synapses with neurons^48^.

As Taylor and Ibrahim eloquently pose^72^, can thought itself drive glioma growth? Venkatesh and colleagues’ seminal work provided evidence of the electrical and synaptic integration of glioma into neural circuit. Our work provides further evidence to the possibility that circuits governing cognition (i.e, MD system) may paradoxically be responsible for glioma growth. By leveraging the Allen Human Brain Atlas, we demonstrate that the MD Core and MD CON networks are both ripe with OPCs. Moreover, gene ontology analyses reveals that MD Core expresses genes related to metabolic processes and signalling pathways and that serotonergic (5HT1b) and dopaminergic (D2) neurotransmission are predominant in the MD Core, while excitatory glutamatergic neurotransmission is specific to the MD CON. Chemotherapy has been shown to deplete OPCs, block activity-dependent myelination, and disrupt cognitive processes. Furthermore, studies on myelin plasticity in both rodents and humans highlight the importance of OPCs in activity-dependent learning. Moreover, evidence from myelin plasticity studies in both rodents and humans suggests the causal importance of this cell-type for activity-dependent learning^27–29^. Ideally, longitudinal functional imaging of glioma patients undergoing the ‘watch-and-wait’ approach would provide an excellent window into If and how gliomas grow and how they perturb functional networks.

### Presurgical Mapping of Hub Architecture

To complement DES and make functional mapping relevant to a neurosurgical context, we aimed to assess the utility of resting-state, or task-negative, functional MRI to capture polysynaptic functional connectivity in glioma patients. We first showed that the MD system is dominated by ‘connector hubs’ – brain regions that occupy a strategic position to serve a critical link between multiple different networks – in healthy individuals across the lifespan. Moreover, resecting five or more connector hubs leads to long-term deficits in glioma patients. This finding was contrasted by the fact that resecting any number of provincial hubs –brain regions that are highly connected within a specific network but have limited connections to other networks –does not lead to long-term cognitive deficits. Thus, this leads to the interpretation that mapping the location of connector hubs can potentially complement surgical planning on where not to operate to avoid long-term cognitive deficits. By removing too many connector hubs from the network, this likely leads to a decline in interhemispheric modularity crucial for maintaining higher-order cognitive functions post-surgery.

The accuracy of non-invasive functional mapping for neurosurgery has been an intense area of investigation^75–78^. Moretto and colleagues recently demonstrated a high-concordance between DES and rsfMRI mapping of brain regions with millimetric precision. While previous studies have explored network-specific connectivity^75^, such as the default mode network, in the context of post-surgical plasticity in glioma patients, no studies have systematically assessed how gliomas integrate within the connectome and how surgery and rehabilitation lead to changes in the modular architecture of the brain^74^. Duffau and others has argued the importance of serial functional imaging to study connectome reorganization in glioma patients^79–80, 89^. Our work provides an initial first step on mapping connector hubs in a neurosurgical context and demonstrates their importance for coordinating brain-wide communication and consequently cognitive tasks.

### Perioperative Modularity for Interventional Neurorehabilitation

Our group previously defined the concept of ‘interventional neurorehabilitation’: connectome-based therapeutic brain stimulation to drive functional recovery after neurosurgery^67^. With this in mind, we focused our analyses on how the unaffected healthy hemisphere in glioma patients is impacted throughout the early surgical period and if we can determine which patients could benefit from interventional neurorehabilitation. Moreover, by avoiding the glioma-affected hemisphere as a brain target, we minimize the risk of eliciting a seizure post-surgery and unknown consequences of electrical stimulation and gliomagenesis. In our CAESAR cohort, we found that decreased perioperative modularity in the healthy hemisphere led to long-term cognitive deficits while maintenance of modularity post-surgery was related to long-term cognitive improvements. From our earlier finding, this finding may be the result of resecting an excess number of connector hubs in the surgical field may directly in decreased interhemispheric connections needed to maintain an optimal modular organisation of the brain for cognitive recovery. Thus, the dissolution and maintenance of the modular organisation of the brain may serve as a valuable biomarker for interventional neurorehabilitation.

Understanding how modularity is related to cognitive outcomes in non-neurosurgical contexts is important to review. Several studies in non-neurosurgical patients point towards baseline modularity indexing cognitive plasticity^26^. A recent review by Gakkeb and D’Epsosito argues that baseline modularity is predictive of intervention-related cognitive plasticity in patients with traumatic brain injury and healthy ageing adults^26^. Specifically, given that modularity is a critical organisational principle of the brain that quantifies the extent to which a network is divided into distinct subnetworks or modules, D’Espositio argues that individuals with higher brain network modularity have larger cognitive gains than those with lower brain network modularity^31^. Baniqued and colleagues demonstrated that baseline modularity is predictive of learning stages in working memory training^80^. The same group demonstrated that older adults with higher baseline connectivity within the associative network had less cognitive decline 3 years later^81^. Thus, it is not surprising that lesions to brain regions important for maintaining global modularity, such as MD regions, leads to more extensive cognitive deficits^82^. Fascinatingly, network modularity increases over the first weeks to months during stroke recovery and is associated with executive function recovery in this patient population^83^. Computational studies also demonstrate that modular networks complete tasks faster and more accurately compared to non-modular network, with associated lower network costs^84^. Finally, modularity is behaviourally relevant – modularity effectively predicts inter-individual variation in working memory capacity and how an individual will respond to various cognitive training^85,86^. Lastly, higher baseline modularity predicts larger cognitive improvements in young healthy adults, during performance of memory tasks^86^, and in children’s scholastic performance^87^. Thus, it is plausible that during cognitive rehabilitation, individuals with higher baseline network modularity have a more optimal network state that requires less network reconfiguration to achieve a successful response to cognitive training.

With regards to practical requirements to translating advanced functional imaging approaches to impacting patient care, it is important that data analysis workflows in neuroimaging has a tremendous scope of flexibility which can affect results^88^. However, given there are multiple methodological approaches to calculating modularity (i.e., number of network nodes, choice of atlas, eyes open/closed during image acquisition, in-scanner head motion), Gallen and D’Esposito found that absolute values of modularity (Q) are not important^33^. In fact, Gallen and D’Esposito found that across studies with methodological differences, baseline modularity and inter-related cognitive gains is robust to varying analytical techniques and is a consistent finding^33^. This suggests that the robustness of this biomarker for low- and high-resolution data and promising in a clinical setting given differences in scanner manufacturers and acquisition times for rsfMRI would be limited to ∼5 minutes given clinical and financial constraints. Thus, based on findings from this Chapter and relevant work done by others, further research is required to replicate this promising finding that modularity may index postsurgical cognitive trajectories in glioma patients. If indeed replicable in independent cohorts, modularity may serve as an effective biomarker to subsequent inform tailored post-surgical interventional rehabilitation

### Limitations and Future Directions

The present study should be interpreted with acknowledging important methodological considerations. Although some analyses were replicated in an independent dataset, there was participant drop-out at multiple time points. Nonetheless, this remains the only deeply sampled, longitudinal dataset that tracks patients through neurosurgical intervention and recovery. In the absence of comparable datasets in the literature, this study serves as a benchmark for future power calculations. Our dataset was limited to frontotemporal tumors, primarily located in associative cortical areas. Although rare, it would be valuable to understand the impact of gliomas in M1, S2, or V1 and their subsequent resection on the connectome. Finally, future studies should determine the cost-benefit ratio of using longer MRI scanning sessions to predict cognitive outcomes post-surgery versus more clinical-grade scanners that are optimized for efficiency rather than in research contexts.

A priority for future studies should be to replicate our analyses of resecting connector hubs on post-surgical long-term cognitive outcomes. Moreover, it is unclear what type of non-invasive brain stimulation protocols are best suited to increase modularity and testing of various sites in the unaffected healthy hemisphere. Lastly, it would be interesting to knw if ‘thought’ or specific functional brain networks can drive gliomagenesis itself. Using this systems neuroscience approach, it would be prudent to determine if gliomas utilize functional networks to proliferate. In that scenario, future studies could be designed to understand which functional networks are preferentially involved in glioma proliferation along with potentially modulating these networks at distant sites to inhibit glioma growth and/or facilitate surgical resection.

## Conclusion

In this work, for the first time, we aimed to develop a systems wide cellular to circuit account of how gliomas and cognition can co-exist. The importance of hub architecture has previously described by but this is the first study to demonstrate the value of functional mapping of hub architecture and how it can practically complement DES. Overall, given that surgical entry zones into the tumour resection cavity are based on anatomical and vascular landmarks, incorporating functional imaging in the preoperative evaluation will likely provide the surgeon additional insight to on the brain’s global network infrastructure governing higher-order cognition. It would be important to determine if we can increase baseline modularity before/after surgery to minimise long-term cognitive deficits in our glioma patients. This can potentially be achieved with neurostimulation and other cognitive enhancement approaches and ushering in ‘interventional neurorehabilitation’. This study paves the way for further investigation into the emerging field of cancer neuroscience.

## Data Availability

All data produced in the present study are available upon reasonable request to the authors

## Acknowledgements

We would like to thank Professor Jennifer Crinion (University College London) and Professor Hugh Markus (University of Cambridge) to provide helpful feedback and comments on the presented work.

